# Functional changes in neural mechanisms underlying post-traumatic stress disorder in World Trade Center responders

**DOI:** 10.1101/2022.04.05.22273447

**Authors:** Azzurra Invernizzi, Elza Rechtman, Paul Curtin, Demetrios M. Papazaharias, Maryam Jalees, Alison C. Pellecchia, Evelyn J. Bromet, Roberto G. Lucchini, Benjamin J. Luft, Sean A. Clouston, Cheuk Y. Tang, Megan K. Horton

**Affiliations:** Department of Environmental Medicine and Public Health, Icahn School of Medicine at Mount Sinai, New York, NY, USA; Department of Radiology and Psychiatry, Icahn School of Medicine at Mount Sinai, New York, NY, USA; Program in Public Health and Department of Family, Population, and Preventive Medicine, Renaissance School of Medicine at Stony Brook University, Stony Brook, NY, USA; World Trade Center Health and Wellness Program, Renaissance School of Medicine at Stony Brook University, Stony Brook, NY, USA; Department of Psychiatry, Renaissance School of Medicine at Stony Brook University, Stony Brook, NY, USA; Department of Environmental Health Sciences, Robert Stempel School of Public Health, Florida International University, Miami, Florida, USA; Department of Medical Surgical Specialties, Radiological Sciences and Public Health, University of Brescia, Italy; Department of Medicine, Renaissance School of Medicine at Stony Brook University, Stony Brook, NY, USA

**Keywords:** resting-state fMRI, graph theory, eigenvector centrality, World Trade Center responders, post-traumatic stress disorder, exposure

## Abstract

World Trade Center (WTC) responders exposed to traumatic and environmental stressors during rescue and recovery efforts have higher prevalence (23%) of persistent, clinically significant WTC-related post-traumatic stress disorder (WTC-PTSD). Here, we applied eigenvector centrality (EC) metrics and data driven methods on resting state functional magnetic resonance (fMRI) outcomes to investigate neural mechanisms underlying WTC-PTSD and to identify how EC shifts in brain areas relate to WTC-exposure and behavioral symptoms. Nine brain areas differed significantly and contributed the most to differentiate functional neuro-profiles between WTC-PTSD and non-PTSD responders. The association between WTC-exposure and EC values differed significantly between WTC-PTSD and non-PTSD in the right anterior parahippocampal gyrus and left amygdala (p= 0.010; p= 0.005, respectively, adjusted for multiple comparisons). Within WTC-PTSD, the index of PTSD symptoms was positively associated with EC values in the right anterior parahippocampal gyrus and brainstem. Our understanding of functional changes in neural mechanisms underlying WTC-related PTSD is key to advance intervention and treatment.

## Main

The men and women involved in the rescue and recovery efforts following the 9/11 World Trade Center (WTC) tragedy were exposed to a complex mixture of smoke, dust and debris generated by the collapse and lasting fires of WTC buildings, as well as traumatic psychosocial stressors including fear for personal safety, injury or illness, exposure to human remains, working long hours and performing arduous work in chaotic conditions^1^. Twenty years after 9/11, WTC responders experience a high prevalence (approximately 23%) of chronic, clinically significant WTC-related posttraumatic stress disorder (PTSD)^2–4^. Recent studies of WTC-responders, including previous studies in our group, leverage structural MRI to characterize anatomical differences between WTC responders with PTSD (WTC-PTSD) and non-PTSD controls ^5,6^ and demonstrate evidence of anatomical changes such as reduced cortical complexity across brain areas (frontal, parietal, and temporal) ^6^. The size of these structural changes was associated with increased PTSD symptoms scales (re-experiencing, avoidance, hyperarousal, negative thoughts) severity^6^. To date, no studies have used functional MRI to characterize PTSD in WTC-responders.

Functional magnetic resonance imaging (fMRI) may prove useful in understanding, detecting and monitoring neurobiological mechanisms underpinning the associations between traumatic psychological and environmental exposures such as 9/11 and PTSD. The spontaneous (task-independent) signal fluctuations observed during resting-state fMRI (rs-fMRI) have been widely used to investigate functional alterations in cortical and subcortical brain areas in psychiatric disorders^7^, while relatively little progress has been made using rs-fMRI to understand underlying mechanisms of PTSD^7,8^. The few existing fMRI and rs-fMRI studies suggest that PTSD follows the ‘fear-conditioning’ paradigm^9–11^ characterized by exaggerated amygdala responses and reduced functionalities in frontal cortical and hippocampal areas^11–14^. Graph theory models leveraging rs-fMRI data provide a comprehensive set of quantitative measures (i.e., centrality, modularity, clustering coefficient, path length) used to investigate global (network-wide) and local (network-specific) aspects of connectivity. Specifically, centrality quantifies the influence of a given brain area on system-wide information flow and integration. Among PTSD subjects, centrality was significantly reduced in hierarchical brain networks.^15–17^ Here, for the first time, we used centrality to better understand neural mechanisms underlying WTC-PTSD and to identify selective shifts in local brain areas that are associated with the WTC-exposure. Identification of local changes in neural activation among WTC responders with PTSD is crucial to advance intervention and may guide treatments protocols such as non-invasive brain stimulation, to modulate brain activity and elicit behavioral changes^18–21^. Furthermore, these selective local changes in PTSD might have disproportionate network effects producing a maladaptive behavior characterized by the set of PTSD specific symptoms including re-experiencing (i.e. involuntary intrusive memories, flashbacks, etc.), avoidance (of distressing thoughts, feelings trauma-related inputs), altered arousal and reactivity (hyperarousal that includes irritability, aggressive and self-destructive behaviors, concentration and sleep problems) and recurrently negative thoughts^7–9^.

In this study, we used eigenvector centrality (EC)^22^ derived from rs-fMRI data^23,24^ together with data-driven methods to: (a) discriminate and identify shifts in centrality between WTC-PTSD and non-PTSD responders; (b) link these shifts to WTC-exposure, and (c) examine how shifts in centrality associate with PTSD symptom scales. We focused on highly connected (or disconnected) nodes (i.e. functional hubs) that influence the global information flow and integration. Based on local EC differences between WTC-responders with and without PTSD, we define functional hubs as key areas to investigate the effect of WTC-exposure on the brain. As an exploratory aim of this study, we examined the association between PTSD symptom scales and the EC values in these functional hubs. This study will expand our knowledge of the biological mechanisms and underlying changes in plasticity of the human brain in WTC-responders that experienced the traumatic exposures at 9/11.

## Results

### Demographic and clinical characteristics

Table 1 reports the clinical and demographic characteristics for the 96 WTC responders included in this study stratified by responders with PTSD (WTC-PTSD) and without PTSD (non-PTSD). Responders were in their mid-fifties at the time of the imaging data acquisition (55.8 ± 5.2 years) and majority males (79%). By design, groups were matched on age at the time of MRI scan, sex, race/ethnicity, and educational attainment. Current major depression diagnosis (MDD), daily use of psychotropic medications, and PTSD symptoms scales (DSM-IV SCID trauma screen) significantly differ between groups. No significant difference in WTC-exposure duration was found between PTSD and non-PTSD WTC responders (average month on site for WTC-PTSD= 3.87 and non-PTSD=4.07, p=0.781). Additional characteristics not included in analysis (i.e., ethnicity, occupation and education level) are reported in supplementary material (Table S1).

**Table 1.** Sociodemographic and clinical characteristics of WTC responders who were selected into the current study (N=96).

| Characteristics | All WTC responders (n=96) | WTC-PTSD (n=45) | non-PTSD (n=51) | p |
| --- | --- | --- | --- | --- |
| **Age (y)** |  |  |  | 0.383 |
| mean ± sd | 55.81 ± 5.26 | 55.31 ± 5.18 | 56.25 ± 5.34 | |
| **Sex (n, %)** |  |  |  | 0.827 |
| Male | 76 (79%) | 37 (49%) | 39 (51%) |  |
| Female | 20 (21%) | 8 (40%) | 12 (60%) |  |
| **MoCA (n)** |  |  |  | 0.736 |
| mean, [range] | 23.23 [12,30] | 23, [12,30] | 23.4 [15,30] |  |
| **Comorbidities (n, %)** |  |  |  |  |
| Major Depressive Disorder |  |  |  | <0.001* |
| No | 78 (81%) | 27 (60%) | 51 (100%) |  |
| Yes | 18 (19%) | 18 (40%) | 0 (0%) |  |
| Cognitive Impairment |  |  |  | 0.424 |
| No | 49 (51%) | 22 (43%) | 27 (53%) |  |
| Yes | 47 (49%) | 23 (45%) | 24 (47%) |  |
| **Medications (n, %)** |  |  |  |  |
| Psychotropic |  |  |  | <0.001* |
| No | 74 (77%) | 27 (60%) | 47 (92%) |  |
| Yes | 22 (23%) | 18 (40%) | 4 (8%) |  |
| Opioid |  |  |  | 0.643 |
| No | 92 (96%) | 43 (95%) | 49 (96%) |  |
| Yes | 4 (4%) | 2 (5%) | 2 (94%) |  |
| **WTC Exposure (mean ± sd)** |  |  |  | 0.781 |
| Total months on site | 3.98 ± 3.31 | 3.87 ± 3.47 | 4.07 ± 3.19 | |
| **DSM-IV SCID Trauma Screen (mean ± sd)** |  |  |  |  |
| Re-experiencing | 17.46 ± 6.74 | 23.4 ± 4.35 | 12.22 ± 3.14 | <0.001* |
| Avoidance | 24.01 ± 9.53 | 32.73 ± 6.12 | 16.31 ± 3.25 | <0.001* |
| Hyperarousal | 17.71 ± 6.67 | 23.97 ± 3.27 | 12.17 ± 2.94 | <0.001* |
| Negative thoughts | 12.19 ± 4.61 | 15.75 ± 4.28 | 9.05 ± 1.66 | <0.001* |

Mean, standard deviation (sd), range (minimum and maximum values), and percentage (%) are reported. P-values quantify differences between WTC-PTSD and non-PTSD were derived using Student *t*-tests for continuous variables and tests for categorical variables. \*denotes significance of $p<0.005$ .

### Centrality differences between WTC-PTSD and non-PTSD

EC values in permutation tests revealed 9 functional hubs where EC values differed significantly between WTC-PTSD and non-PTSD groups; right and left anterior inferior temporal gyrus, right superior parietal lobule, right anterior parahippocampal gyrus, right anterior and posterior temporal fusiform cortex, right caudate nucleus, left amygdala and the brainstem (Table 2, Figure 1). PLS-DA analysis of EC values indicates a sufficient signature to separate WTC-responders with and without PTSD (Figure 2A). The ROC illustrating the sensitivity and specificity for predicting WTC-PTSD from the first discriminant axis yields an AUC of 0.749 (0.651-0.847; Figure 2B). Figure 3 shows the loading of each brain region (n=111) on a given dimensional subscales to determine which regions contribute most to the overall divergence between WTC responders with and without PTSD based on the PLS-DA analysis. The same 9 functional hubs identified in permutation tests had the highest loadings in the PLS-DA, therefore contributing most to differentiate functional neuro-profiles of WTC responders with and without PTSD (Figure 3). Notably, of the 9 functional hubs, 7 were localized in the right hemisphere.

**Table 2.** Statistical differences in eigenvector centrality between WTC-PTSD and non-PTSD.

| WTC-PTSD versus non-PTSD |  |  |  |
| --- | --- | --- | --- |
| Brain region (ROIs) | Hemisphere | Abbreviation | p |
| Inferior Temporal Gyrus (anterior) | R | ITG | 0.042 |
| Superior Parietal Lobule | R | SPG | 0.018 |
| Parahippocampal Gyrus (anterior) | R | PHG | 0.048 |
| Temporal Fusiform Cortex (anterior) | R | FFG | 0.012 |
| Temporal Fusiform Cortex (posterior) | R | STG | 0.017 |
| Caudate nucleus | R | CAU | 0.015 |
| Brain Stem | - | - | 0.016 |
| Inferior Temporal Gyrus (anterior) | L | ITG | 0.015 |
| Amygdala | L | AMYG | 0.014 |

**Figure 1.**
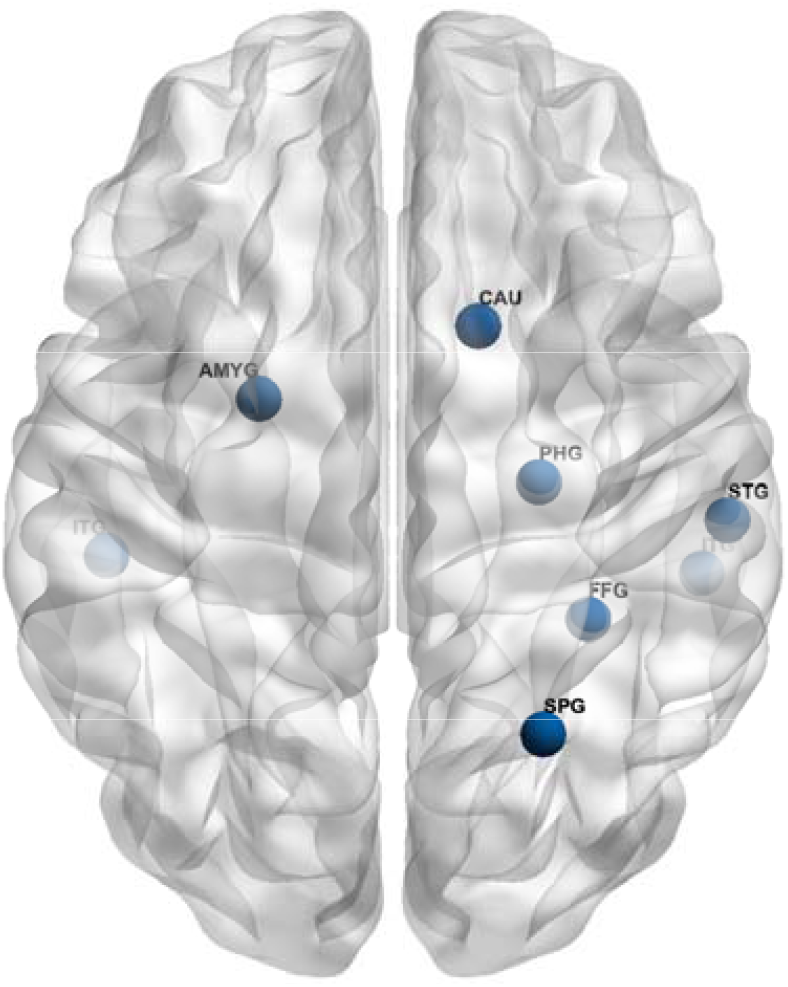
Functional hubs differ in eigenvector centrality between WTC-PTSD and non-PTSD.

**Figure 2.**
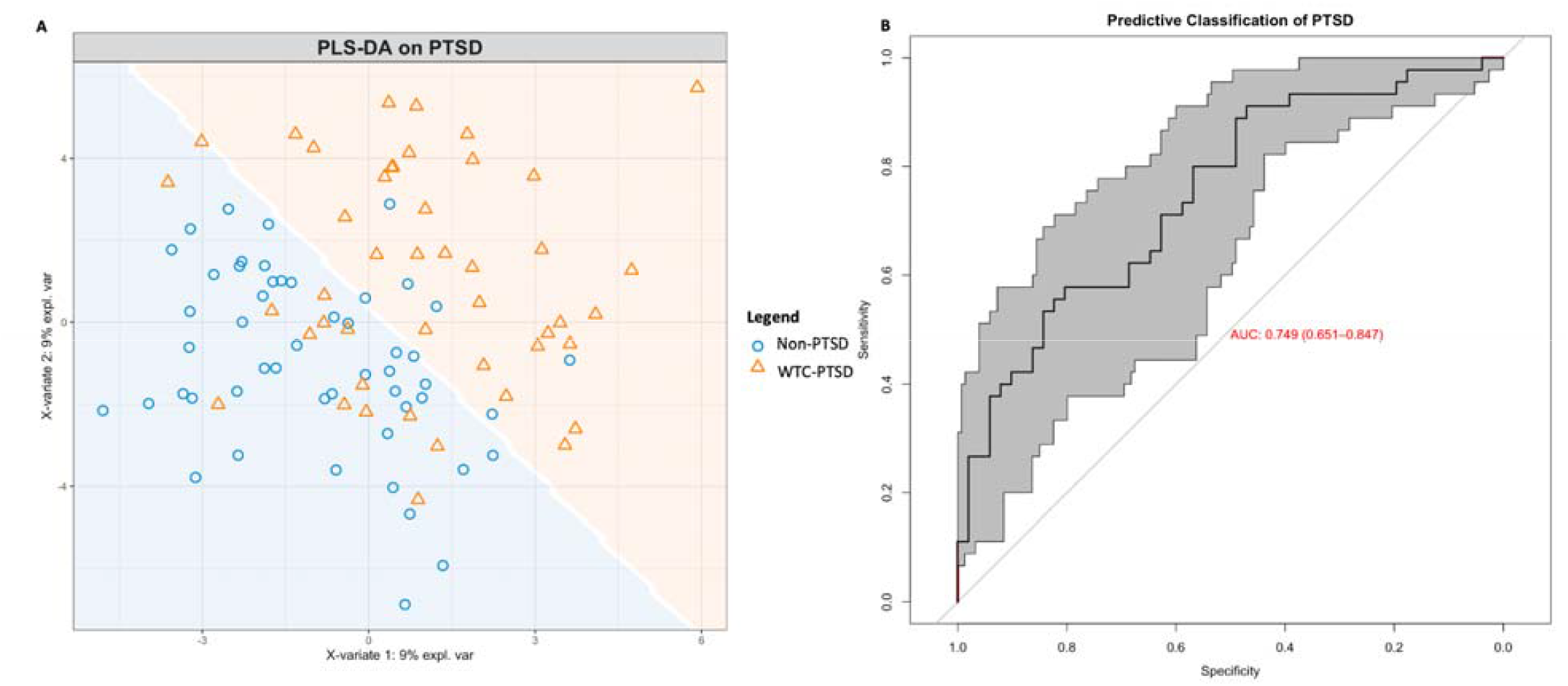
PLS-DA signature separating eigenvector centrality values between WTC-PTSD and non-PTSD groups. Using the eigenvector centrality (EC) values from 111 brain regions as inputs, the partial least squares discriminant analysis (PLS-DA) constructs a lower-dimensional subspace to maximize the separation between WTC-responders with and without PTSD. *Panel A* - Orange triangle (WTC-PTSD) and blue dots (non-PTSD) show individual participants scores on the first (x-axis) and second (y-axis) discriminate axes derived through PLS-DA. Separation of orange triangle and blue dots (white line) illustrates the prediction area associated with each class, with subjects scoring in orange space predicted to be WTC-responders with PTSD, while those in blue space are predicted to be WTC-responders without PTSD. *Panel B* - Receiver operating characteristic (ROC) curve illustrating the sensitivity and specificity for predicting WTC-PTSD from the first discriminant axis with varying classification thresholds. To quantify uncertainty in the ROC curve and discriminant prediction, a confidence interval of the ROC curve was computed using 2000 stratified bootstraps, yielding an area-under-the-curve (AUC) of 0.749 (0.651-0.847).

**Figure 3.**
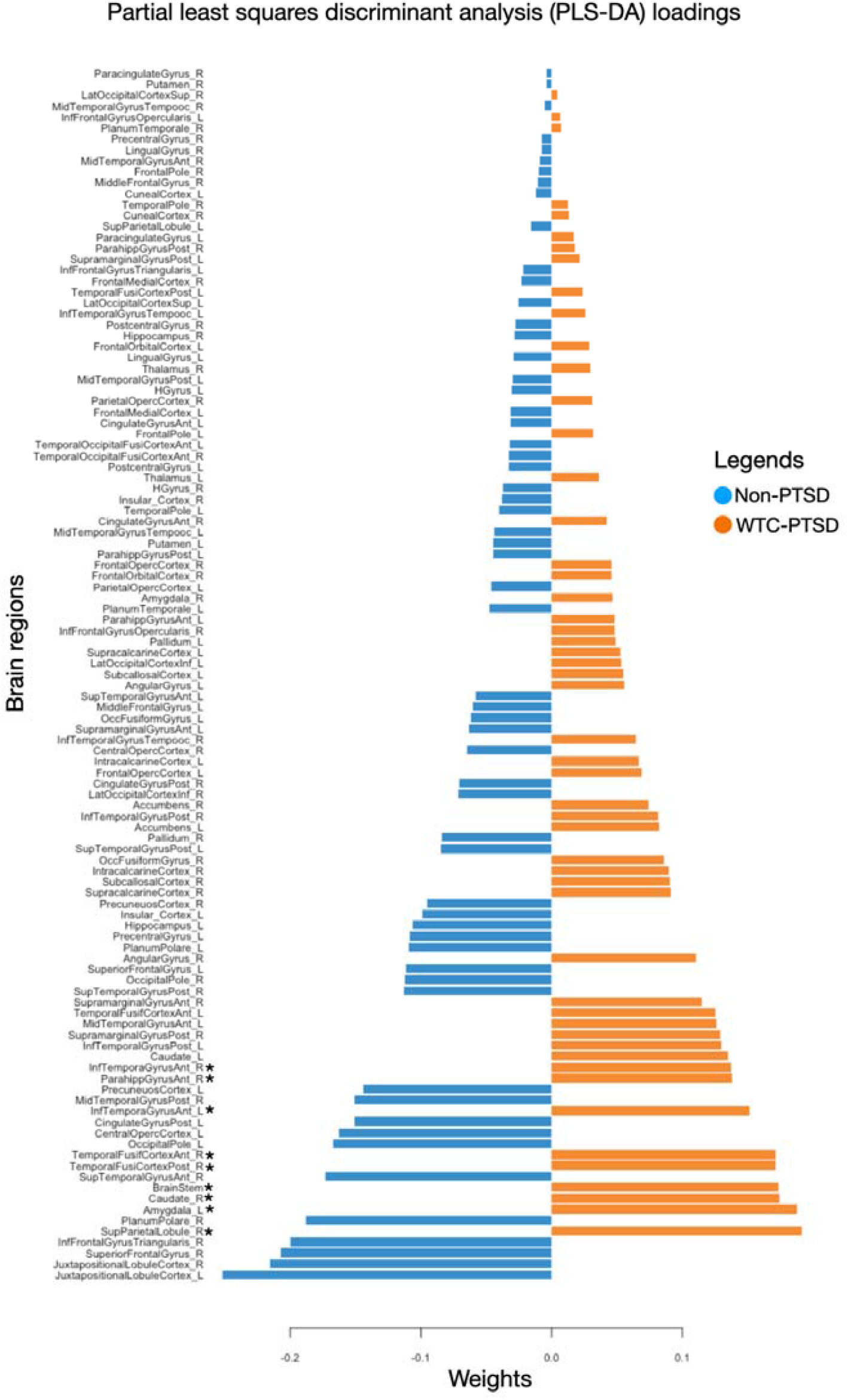
Partial least squares discriminant analysis (PLS-DA) loadings contributing to the divergence of WTC-PTSD from non-PTSD. PLS-DA identifies the loading of each brain region (n=111) on a given dimensional subscale to determine which regions contribute most to the overall divergence between WTC responders with and without PTSD. Loadings are color coded to indicate the group with a higher mean score on that variable, blue lines indicate variable loadings non-PTSD, and orange lines indicate variable loadings in WTC-PTSD. The loading direction (positive or negative) indicates the direction of the associations between individual brain regions and PTSD. * indicates p < 0.05 in brain areas where EC values differ significantly between groups.

### Centrality and WTC Exposure

Results from the GLM suggest that WTC exposure duration (months on site) moderates the association between PTSD and EC values in two of the 9 functional hubs; the right anterior parahippocampal gyrus and the left amygdala (p for interaction = 0.010 and 0.005, respectively adjusted for multiple comparisons; Table 3, Figure 4). In WTC-PTSD responders, prolonged WTC exposure is associated with decreased EC values in these two brain areas, (p=0.05 and 0.002, 95% CI [-0.0005, 0.001; -0.0003, -0.0006], respectively). For completeness, remaining hubs and models uncorrected for FWE are reported in supplementary materials (Table S2, Figure S1, Table S3).

**Table 3.** Association between EC values, PTSD status and WTC exposure duration.

| Parahippocampal Gyrus (anterior, right) |  |  |  | Amygdala (left) |  |  |  |
| --- | --- | --- | --- | --- | --- | --- | --- |
| Predictors | Estimates | CI | $p$ | Predictors | Estimates | CI | $p$ |
| Intercept | 0.084 | 0.082 - 0.087 | **<0.001** | Intercept | 0.086 | 0.084 - 0.088 | **<0.001** |
| Months on site | 0.000 | -0.000 - 0.001 | 0.099 | Months on site | 0.000 | -0.000 - 0.001 | 0.492 |
| Psychotropic | -0.002 | -0.005 - 0.002 | 0.307 | Psychotropic | -0.002 | -0.005 - 0.000 | 0.063 |
| Opioid | -0.002 | -0.008 - 0.005 | 0.576 | Opioid | -0.008 | -0.014 - -0.003 | **0.001** |
| MDD | -0.002 | -0.005 - 0.002 | 0.367 | MDD | 0.002 | -0.001 - 0.005 | 0.109 |
| PTSD | 0.007 | 0.003 - 0.011 | **<0.001** | PTSD | 0.005 | 0.002 - 0.008 | **0.001** |
| Months on site*PTSD | -0.001 | -0.002 - 0.000 | **0.01** | Months on site*PTSD | -0.001 | -0.001 - -0.000 | **0.005** |
| Observations | 86 |  |  | 86 |  |  |  |
| R2 | 0.162 |  |  | 0.332 |  |  |  |

**Figure 4.**
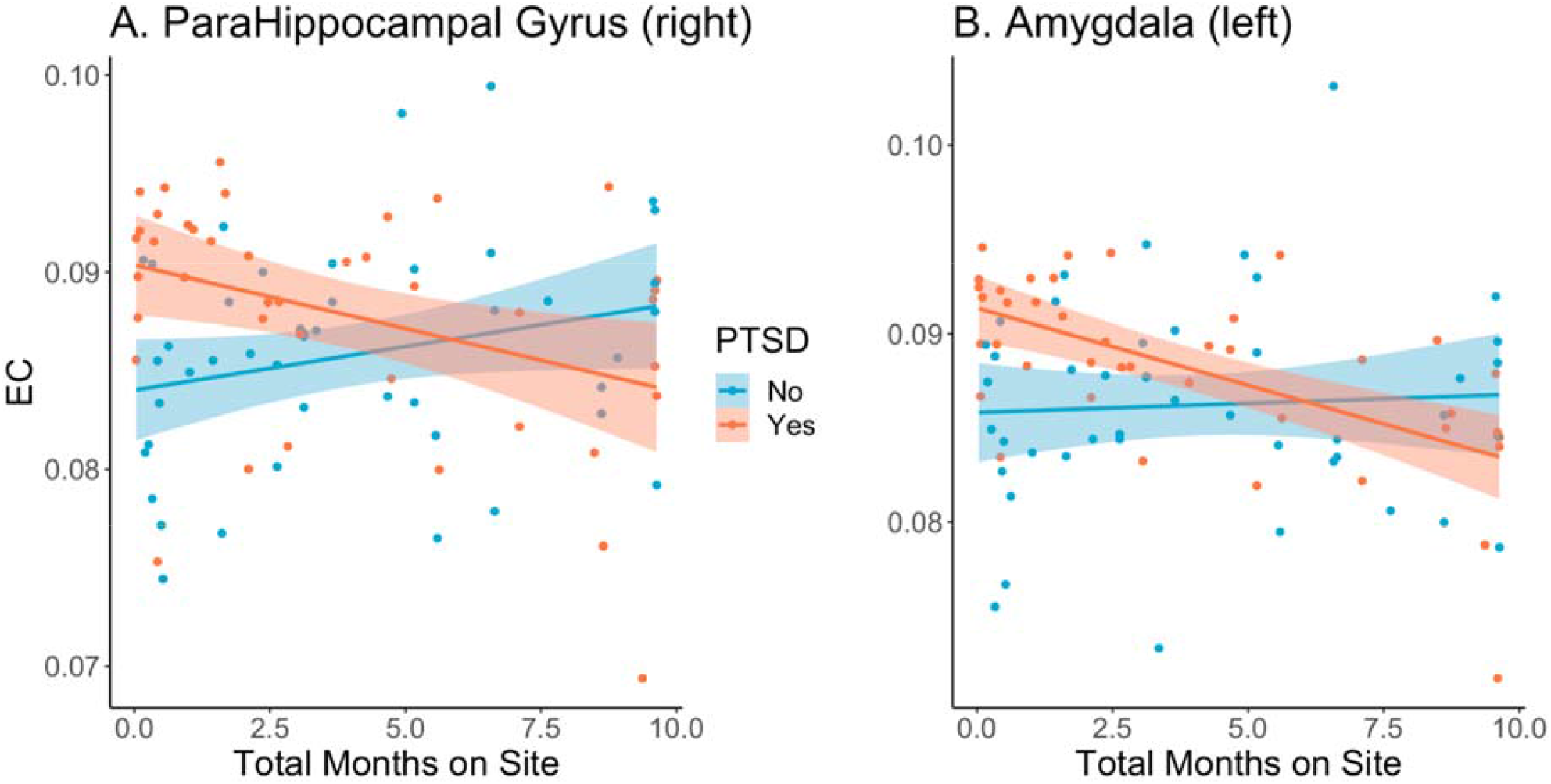
WTC exposure duration and EC values. These graphs plot the relationship (interaction) between WTC exposure duration in months (x-axis) and eigenvector (EC) values (y-axis) stratified by WTC-PTSD (orange dots) and non-PTSD (blue dots) for the right anterior parahippocampal gyrus (A) and the left amygdala (B). WTC exposure duration (months on site)moderates the association between PTSD status and EC values in both hubs (p_Parahippocampal Gyrus_= 0.010 and p_Amygdala_= 0.005).

### Centrality and PTSD symptoms

Within the WTC-PTSD group, we identified significant associations between EC values and the weighted PTSD symptom index in the right anterior parahippocampal gyrus (β=-0.002, SE=0.0009, p=0.048; Table 4) and the brainstem (β=-0.001, SE=0.0007, p=0.046; Table 4). Avoidance symptoms contributed 64.3% to the overall association between EC values and PTSD symptoms index in the right anterior parahippocampal gyrus (Figure 5, panel A). Hyperarousal contributed 65.4% to this association in the brainstem (Figure 5, panel B). However, none of these associations survived correction for multiple comparisons.

**Figure 5.**
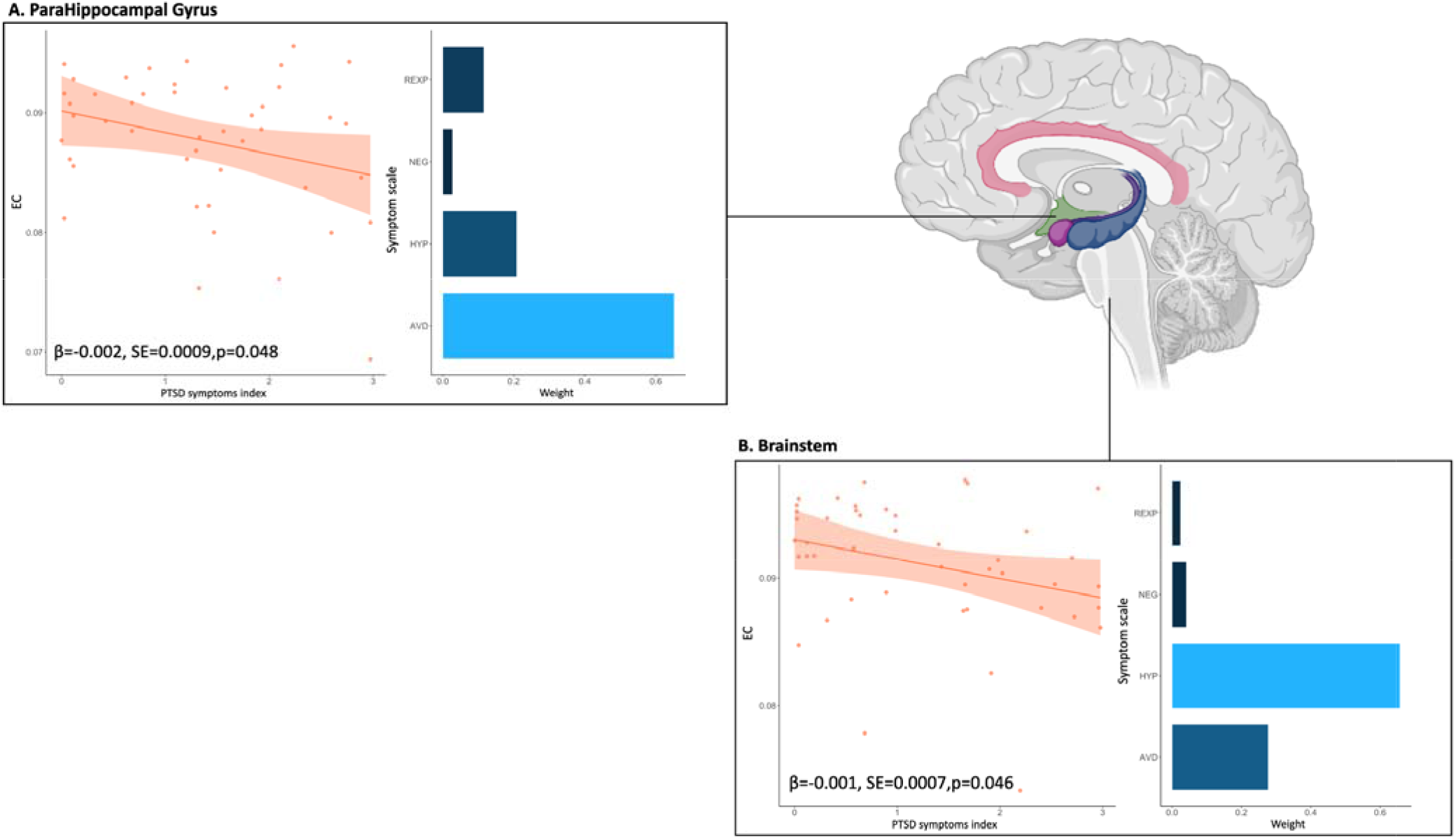
PTSD-symptom scales and centrality. WQS regression analysis of the association between weighted index of PTSD symptoms and EC values for the right anterior parahippocampal gyrus and the brainstem (panel A and B, respectively) .t. EC values of the right and left anterior inferior temporal gyrus have been averaged into one unique ROI (anterior inferior temporal gyrus) for this analysis. In the graph, orange dots represent WTC-PTSD responders, the orange line represents the association between the PTSD symptoms index and EC values, shaded orange is the confidence interval. The histogram displays the contribution of each PTSD symptom to this association and each bar represents a different symptom scale. Symptoms with the highest weights are plotted in light blue.

**Table 4.**
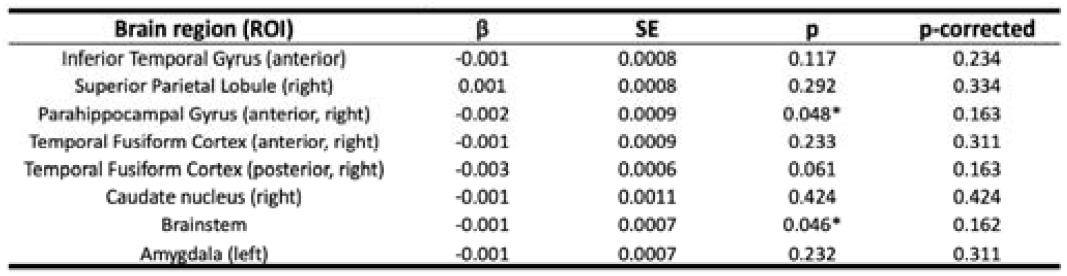
PTSD-symptom scales and centrality. WQS regression analysis of the association between weighted index of PTSD symptoms and EC values for the right anterior parahippocampal gyrus and the brainstem (panel A and B, respectively) .t. EC values of the right and left anterior inferior temporal gyrus have been averaged into one unique ROI (anterior inferior temporal gyrus) for this analysis. In the graph, orange dots represent WTC-PTSD responders, the orange line represents the association between the PTSD symptoms index and EC values, shaded orange is the confidence interval. The histogram displays the contribution of each PTSD symptom to this association and each bar represents a different symptom scale. Symptoms with the highest weights are plotted in light blue.

**Figure 5 and Table 4.**
| Brain region (ROI) | $\beta$ | SE | p | p-corrected |
| --- | --- | --- | --- | --- |
| Inferior Temporal Gyrus (anterior) | -0.001 | 0.0008 | 0.117 | 0.234 |
| Superior Parietal Lobule (right) | 0.001 | 0.0008 | 0.292 | 0.334 |
| Parahippocampal Gyrus (anterior, right) | -0.002 | 0.0009 | 0.048* | 0.163 |
| Temporal Fusiform Cortex (anterior, right) | -0.001 | 0.0009 | 0.233 | 0.311 |
| Temporal Fusiform Cortex (posterior, right) | -0.003 | 0.0006 | 0.061 | 0.163 |
| Caudate nucleus (right) | -0.001 | 0.0011 | 0.424 | 0.424 |
| Brainstem | -0.001 | 0.0007 | 0.046* | 0.162 |
| Amygdala (left) | -0.001 | 0.0007 | 0.232 | 0.311 |

## Discussion

This is the first ever study to use rs-fMRI to investigate local connectivity differences between PTSD and non-PTSD WTC responders. Using graph-based network metrics and data-driven methods, we identified clear differences in functional neuro-profiles of WTC-responders with and without PTSD in nine brain functional hubs. PTSD status modified the association between duration of WTC exposure (i.e., months at the rescue and recovery site) and shifts in EC. This was observed in two functional hubs known to be associated with PTSD; the right anterior parahippocampal gyrus and the left amygdala. Finally, within the WTC-PTSD group, we observed associations between EC values and PTSD symptoms in the right anterior parahippocampal gyrus and in the brainstem. Our results confirmed previous results presented in literature, and furthers contribute to our understanding of the neurobiological underpinnings of WTC-related PTSD. These results may guide treatment efforts and inform future disaster-response activities.

Seven out of nine functional hubs differ between WTC-PTSD and non-PTSD in our study were located in the right hemisphere (Figure 1, Table 2), suggesting lateralization of the association between WTC-PTSD and centrality. These results align with our previous study in this cohort demonstrating anatomical changes with strong lateralization in the right hemisphere across several brain areas^6^. Lateralization may be due to specific functions of the right hemisphere such as the continuous interchange of information with the amygdala. The amygdala plays a key role in in PTSD^25–27^ and is involved in personality, emotional, and behavioral regulation^28,29^, fear and fear conditioning^30,31^, and memory of stressful events^32^. Notably, the left hemisphere that is mostly involved with verbal communication and problem-solving abilities, seems to be less associated with PTSD in our study and in others ^6,33^. Taken together, these results of lateralizatized changes for both functional and structural data in WTC-responders^6^ align with previous studies performed in traumatized subjects^33–35^, where the right hemisphere appears to be more generally affected by PTSD^6,33^ when compared to the left hemisphere.

Duration of WTC exposure (i.e., number of months spent at the WTC site in rescue and recovery efforts) did not differ between responders with PTSD and those without PTSD (Table 1). However, among responders with PTSD, decreased EC values in the right anterior parahippocampal gyrus and the left amygdala, are associated with prolonged WTC exposure to traumatic events during search and rescue efforts at and for months after 9/11 (Figure 4, Table 3). During these months on the pile, WTC responders were exposed to both traumatic event and a number of pollutants including lead, polycyclic aromatic hydrocarbons (PAHs), polychlorinated biphenyls (PCBs), and dioxins that all together might have played a role in the anatomical and functional changes observed^36–39^. These changes and their significant association with long-term WTC exposure involve only specific cortical and subcortical brain regions such as the whole hippocampus and its subfields^5^, parahippocampal gyrus, amygdala, and finally frontal and parietal brain regions^6^ that seem more vulnerable to experience at the WTC site. These results might be explained by the fear-conditioning mechanism and novel neurocircuitry models^9–11^ that suggest the triggering event, in this case WTC responders experienced during rescue and recovery efforts, targets brain areas known to be involved in PTSD, i.e., the parahippocampus and amygdala. Our results are consistent with previous studies on the WTC cohort that found associations between longer-term WTC exposure and structural changes defined as reduced cortical volumetric and cortical complexity^5,6^.

PTSD is characterized by recurrent, intercorrelated symptoms such as re-experiencing, avoidance, negative affect, and hyperarousal. Neurobiological models of PTSD show that each of these symptoms are associated with changes in specific brain areas^8^. Disentangling the unique contribution of each PTSD symptom within the centrality neuroprofiles in the WTC-PTSD group contributes to our understanding of neurobiological mechanisms underpinning WTC related PTSD. In our cohort, avoidance symptoms contributed most to the association between overall PTSD symptoms cores and EC shifts in the parahippocampal gyrus. The parahippocampal gyrus surrounds the hippocampus and is part of the temporal lobe network. Parahippocampal gyrus function is crucial for encoding and retrieving episodic, spatial and contextual memories ^40–44^. Consistent with these functions, previous studies linked this cortical brain area with avoidance behavior, disrupted encoding of episodic and autobiographical memories and functional changes in PTSD subjects^41,45^. Hyperarousal symptom contributed the most to the association between PTSD symptoms scores and and centrality values (Figure 5). The brainstem is critical to generate and maintain the general arousal state and to provide the trigger for innate, reflexive defensive responses^46,47^. Previous studies showed that prolonged and repeated traumatic experiences lead to changes in this brain area and emerging evidence suggests its critical role in the neurobiological model of PTSD^7,48,49^.

Taken together, our findings show functional changes and association with WTC-exposure in the brainstem, amygdala, and the parahippocampal gyrus areas in WTC-PTSD responders. Similar to findings from a previous study in these WTC responders, we did not find significant changes in the hippocampus^5^. Our findings are further consistent with several studies demonstrating the relevance of these cortical and subcortical areas in the innate threat processing-related network, well-connected brain areas responsible for triggering the alert and defense mechanisms by a fast communication between deep-to higher-layer of brain regions, in PTSD and in its subtype^50–52^. Unbalanced and disrupted patterns of communication between the amygdala and parahippocampal gyrus have been widely reported in PTSD^7,53^. In agreement with these studies^7,53^, we report an exaggerated influence of the amygdala on system-wide integration in WTC-PTSD. Finally, our understanding of the neural mechanisms underlying WTC-PTSD is crucial for the progression of novel treatments and interventions that are still lacking in this disorder^8,20,21^. Treatments based on transcranial magnetic stimulation and deep brain stimulation might consider the use of rs-fMRI data together with targeting data-driven methods (i.e. graph theory) to customize the intervention by modulating relevant neural networks^8,21,24^.

The 9/11 rescue and recovery workers were exposed to a mixture of environmental and traumatic psychosocial stressors that allow us to learn more about the impact of extreme, prolonged exposure on the brain. In this study for the first time, we identified key brain areas associated with WTC-exposure that may guide treatment efforts and inform future disaster-response activities. Further, our data-driven approach could be used to track emerging conditions following future man-made and natural disasters. Limitations of our study design include the small sample size and lack of external control group (non-WTC). Our small sample size prohibited splitting the sample into training and validation subset. A larger sample size may also provide more statistical power which could identify additional functional hubs associated with WTC-PTSD. In addition, a strong effort was made to increase the recruitment of underrepresented populations including women and people of color to the point of doubling the numbers of both in this sample compared to the responder population enrolled in our program ^54–56^, our sample would benefit from improved diversity. The EAQ, from which we gather self-reported experience during WTC rescue and recovery efforts, was administered at any time following 9/11 (ranging from 1 year to 20 years) and may be subject to recall bias. Finally, we lack accurate assessments of life trauma and/or PTSD status and MRI scans in WTC responders prior to 9/11, and we lack a comparison group of responders with subsyndromal, mild, heterogeneous, or remitted PTSD. All together, these limitations prevent us from generalizing our findings to PTSD in the population.

To conclude, this is the first ever study using rs-fMRI data to provide novel insights into the underlying neural mechanisms and changes in plasticity of the human brain in WTC-responders that experienced the traumatic exposures at 9/11. Our results suggest that responders who developed WTC-related PTSD present significant brain functional changes in specific brain areas previously shown to be associated with PTSD. These changes are associated with WTC-exposure, as well as with PTSD symptomatology.Future studies to elucidate the different contributing factors to the etiology of PTSD in WTC responders are still needed to advance our understanding in this debilitating disease and to help intervention and treatment.

## Methods

### Participants

Ninety-nine participants were recruited from a single clinic-based monitoring WTC Health program who previously participated in an epidemiology study of cognitive accelerated aging. Complete details of the study can be found elsewhere^54,57–59^. Briefly, all participants were aged 44 to 65 years, fluent in English and satisfied eligibility criteria for MRI scanning (i.e., metal implants or shrapnel, claustrophobia, no prior history of traumatic brain injury, body mass index (BMI) ≤ 40). Up to 8 weeks before MRI, global cognitive status was objectively assessed using the Montreal Cognitive Assessment (MoCA)^57^. The diagnostic assessment of PTSD related to WTC (WTC-PTSD) was determined from a structured diagnostic interview, described in detail below. Upon enrollment, WTC-PTSD case and non-PTSD control groups were matched on age within 5 years, sex, race/ethnicity and type of responders (i.e., police)^54,60^. From the 99 participants who completed the MRI scan, 3 subjects were excluded for poor quality MRI data (i.e., excessive movement, reduced field), leaving 96 participants included in this project. Study procedures that follow the Declaration of Helsinki, were approved by the Institutional Review Boards at both Stony Brook University and the Icahn School of Medicine at Mount Sinai. Informed consents were signed by all participants at enrollment after all study procedures were fully explained.

### Neuropsychological assessment

PTSD diagnosis was assessed using the Structured Clinical Interview for the DSM-IV (SCID-IV)^61^, a semi-structured interview schedule administered by trained clinical interviewers. The PTSD module used WTC exposures as the index trauma. In particular, SCID items were modified to assess PTSD symptoms in relation to traumatic WTC exposure events (i.e. the worst episode of symptoms since 11 September 2001)^60^. Eligibility criterion for WTC-PTSD status was presence/absence of current PTSD diagnosis at the time of enrollment into the current study. WTC-PTSD status was considered ‘present’ if criteria were reported in the past 3 months at the time of the interview^60^. PTSD symptom subdomains were measured using continuous subscales calculated using reported symptom severity in the SCID-IV for the following symptom domains: re-experiencing, avoidance, hyperarousal, and negative thoughts symptoms (scores ranging from [10-30], [14-42], [10-30], [8-24], respectively). Major depressive disorder (MDD) was assessed using the SCID-IV and the presence or absence of current (i.e., active in the past month) MDD diagnosis was determined. MDD was not an exclusion criterion.

### WTC exposure duration

An interviewer-administered exposure assessment questionnaire (EAQ) was administered to all WTC-responders upon enrollment in the CDC/NIOSH supported WTC General Responders Cohort and collected at the first monitoring visit of the epidemiologic study only^58^. The EAQ served to assess exposure to potentially harmful physical and psychological conditions from participation in the WTC related rescue and recovery efforts (i.e. use of personal protective equipment at the WTC site). In this study, WTC exposure was calculated as time spent (months)of working on the WTC site^55,60^. WTC exposure (i.e., #months duration at the WTC site, ranged from 0-10 months). This exposure variable was not available for 10 participants, therefore analyzes including this variable were done using a sample of 86 WTC responders. There is no significant difference in demographic characteristics or PTSD status between the participants removed (n=10) and the participants included in the analysis (n=86).

### MRI and fMRI data acquisition

Magnetic resonance imaging (MRI) and functional MRI (fMRI) data acquisition was performed on a high-resolution 3-Tesla SIEMENS mMR Biograph scanner using a 20 channel head and neck coil. For each WTC responder, a high-resolution 3D T1-weighted structural scan was acquired using a MPRAGE sequence (TR =1900ms, TE= 2.49ms, TI=900ms, flip angle =9, acquisition matrix=256×256 and 224 slices with final voxel size=0.89×0.89×0.89 mm). Then, functional T2*-weighted, 2D echo planar images were obtained using the following parameters: TR= 1500 ms, TE=27 ms, pulse angle=80 degrees, field of view=22 cm, with a 74 × 74-pixel matrix and a slice thickness of 2.5 mm. Fifty contiguous oblique-axial sections to cover the whole brain where the first four images were discarded to allow the magnetization to reach equilibrium. For each subject, a single 10-minute continuous functional sequence was acquired, for a total of 400 volumes. During resting-state scan, participants were instructed to relax, lay awake, not think about anything, lie still, with their eyes open. Padding was used for a balance between comfort and reduction of head motion.

### Data availability

De-identified data will be made available upon reasonable request to the corresponding author; raw image files can be accessed upon completion of a data use agreement.

### fMRI data analyses

Image pre-processing, eigenvector centrality mapping (ECM), and statistical analyses were performed using SPM12 (Wellcome Department of Imaging Neuroscience, London, UK), fastECM toolbox^22^ and customized scripts, implemented in MatLab 2016b (The Mathworks Inc., Natick, Massachusetts) and R (v3.4).

#### Image preprocessing

For each subject, the structural magnetic resonance image was co-registered and normalized against the Montreal Neurological Institute (MNI) template and segmented to obtain white matter (WM), gray matter (GM) and cerebrospinal fluid (CSF) probability maps in the MNI space. FMRI data were spatially realigned, co-registered to the MNI-152 EPI template and subsequently normalized utilizing the segmentation option for EPI images in SPM12. All normalized data were denoised using ICA-AROMA^62^. Additionally, spatial smoothing was applied (8 millimeters) to the fMRI data. No global signal regression was applied.

Based on the Harvard-Oxford^63^ atlas, 111 regions of interest (ROI; 48 left and 48 right cortical areas; 7 left and 7 right subcortical regions and 1 brainstem) were defined. For each ROI, a time-series was extracted by averaging across voxels per time point.

Then, to facilitate statistical inference, data were “pre-whitened” by removing the estimated autocorrelation structure in a two-step GLM procedure^64,65^. In the first step, the raw data were filtered against the 6 motion parameters (3 translations and 3 rotations). Using the resulting residuals, the autocorrelation structures present in the data were estimated using an Auto-Regressive model of order 1 (AR(1)) and then removed from the raw data. Next, the realignment parameters, white matter (WM) and cerebrospinal fluid (CSF) signals were removed as confounders on the whitened data.

#### Eigenvector Centrality Mapping (ECM)

Fast ECM^22^ was performed on the defined ROI time course data per subject. The ECM method builds on the concept of node centrality, which characterizes functional networks active over time and attributes a voxel-wise centrality value to each ROI. Such a value is strictly dependent on the sum of centrality properties of the direct neighbor nodes within a functional network. In the fast ECM toolbox^22^, ECM is estimated from the adjacency matrix, which contains the pairwise correlation between the ROIs. To obtain a real-valued EC value, we added +1 to the values in the adjacency matrix. Several EC values can be attributed to a given node, but only the eigenvector with the highest eigenvalue (EV) will be used for further analyses. The highest EV values were averaged across subjects at group level. Based on these values, influential ROIs, i.e. the hubs ^66–68^, can then be identified.

### Statistical analyses

#### Descriptive statistics

By study design, sample subgroups (WTC-PTSD and non-PTSD) were matched for age at the time of the visit, sex, race/etchnicity and education level^2,6,58,60^. Pairwise Student t-tests with Welch’s correction for continuous variables and x_z_ tests for categorical variables were used to examine differences in clinical and demographic characteristics across the groups.

#### Permutation statistics

We quantify possible functional hubs (i.e., brain regions with EC values that differ significantly between WTC-PTSD and non-PTSD) by permuting labels with 10000-time repetitions. ROIs with statistically significant (p≤ 0.05) EC values between WTC-PTSD and non-PTSD were considered functional hubs. Family-wise error (FWE) correction was applied for the number of group level comparisons, not for the total number of ROIs analyzed. Only p-values FWE corrected and adjusted for use of medications (psychotropic and opioids) and current depression (MDD) are reported.

#### Partial least squares discriminant analysis

To model the divergence of EC values between WTC responders with and without PTSD, we implemented partial least squares discriminant analysis (PLS-DA)^69,70^. Using the EC values from 111 ROIs as inputs, this supervised dimensionality-reduction technique constructs a lower-dimensional subspace that maximizes the separation between WTC responders with and without PTSD. In contrast to traditional linear modeling strategies, which are evaluated on the basis of statistical significance, the utility of this approach is evaluated on the basis of predictive efficacy; that is, how well do derived dimensions discriminate between WTC responders with and without PTSD. A receiver operating characteristic (ROC) curve was generated to predict WTC-PTSD from the first discriminant axis with varying classification thresholds^71^. To quantify uncertainty in the ROC curve and discriminant prediction, a confidence interval of the ROC curve was computed using 2000 stratified bootstraps. Furthermore, we implemented a rank-based statistical test to determine if the performance of our classification algorithm was significantly better than a classification made randomly. P-values derived from this method should be interpreted in the context of the model’s overall predictive performance, rather than indicative of any individual feature. Finally, the loadings of each region on a given dimensional subscale were evaluated to determine which regions contributed most to the overall divergence between WTC responders with and without PTSD. PLS-DA models were implemented in R using the mixOmics package^72^.

#### General Linear Model

To test our hypothesis that WTC exposure duration (months on site) moderate the association between PTSD and EC values in functional hubs, general linear model (GLM) regressions were computed using current PTSD diagnosis and cumulative WTC exposure duration expressed in months as predictors. These models were adjusted for the use of medications (psychotropic and opioids) and current depression diagnosis. Only EC values of brain areas identified as hubs were entered as outcomes in this analysis. GLMs were implemented using R (Version 1.4.1717).

#### Generalized Weighted Quantile Sum Regression

To determine associations between EC in the functional hubs and PTSD symptoms, we used weighted quantile sum (WQS) regression^73^. WQS is a data driven, mixtures-based ensemble modeling strategy that tests for associations between the combined effect of multiple, correlated variables and an outcome of interest. Here, we include four dimensional scales of PTSD as predictors: re-experiencing, avoidance, hyperarousal, and negative thoughts. The WQS analysis is implemented in two steps. First, a weighted index representing the association between each individual dimensional scales of PTSD and EC was estimated across 5000 bootstrap samples. Second, this weighted index was tested in a linear regression model predicting the association between the “mixture” of the PTSD scales and EC. Prior to model estimation, all exposures were quartiles. The mixture of PTSD symptom scale is defined such that 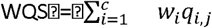 is the sum of the cross products of the empirically estimated weight (w_i_) for each predictor variable (i) and the ranked concentration of that predictor per subject (q_i,j_). A significance test for the WQS index provides an estimate of the association with the overall PTSD symptom scales, while the weights associated with each predictor provide an indicator of each individual variable’s contribution to the overall effect. All weights are constrained to sum to one, enabling sorting by relative importance. Factors that impact the outcome have larger weights; factors with little or no impact on the outcome have near-zero weights. Only WTC-PTSD responders were included in this analysis. These models were adjusted for use of medications (psychotropic and opioids) and current depression (MDD).

## Data Availability

All data produced in the present study are available upon reasonable request to the authors

## Acknowledgements

The authors would like to acknowledge support from the Centers for Disease Control and Prevention for supporting the neuroimaging study (CDC/NIOSH U01 OH011314), the National Institute on Aging that supports research on characterization and treatment of Alzheimer’s disease (NIH/NIA P50 AG005138), and aging-relates work in this population (NIH/NIA R01 AG049953). We would also like to acknowledge ongoing funding to monitor World Trade Center responders as part of the WTC Health and Wellness Program (CDC 200-2011-39361).

## Supplementary material

**Table S1.** Sociodemographic characteristics of WTC responders who were selected into the current study (N=96). Mean, standard deviation (sd), range (minimum and maximum values), and percentage (%) are reported. P-values quantify differences between WTC-PTSD groups and the noted characteristics; they were derived using Student *t*-tests for continuous variables and tests for categorical variables. *denotes significance of *p*<0.005.

| Characteristics | All WTC responders (n=96) | PTSD- (n=51) | PTSD+ (n=45) | p |
| --- | --- | --- | --- | --- |
| **Ethnicity (n,%)** |  |  |  | 0.804 |
| White | 73 (76.04%) | 38 (74.51%) | 35 (77.7%) |  |
| Black | 10 (10.42%) | 6 (11.76%) | 4 (8.88%) |  |
| Other | 13 (13.54%) | 7 (13.72%) | 6 (13.33%) |  |
| **Occupation (n,%)** |  |  |  | 0.725 |
| NYPD | 56 ( 58.33%) | 35 (68.63%) | 21 (46.66%) |  |
| Other | 38 (39.58%) | 15 (29.41%) | 23 (51.1%) |  |
| **Education (n,%)** |  |  |  | 0.034 |
| High school or less | 22 (22.92%) | 9 (17.65%) | 13 (28.88%) |  |
| College | 31 (32.29%) | 14 (27.45%) | 17 (37.78%) |  |
| University degree | 43 (44.79%) | 28 (54.90%) | 15 (33.33%) |  |

**Figure S1.**
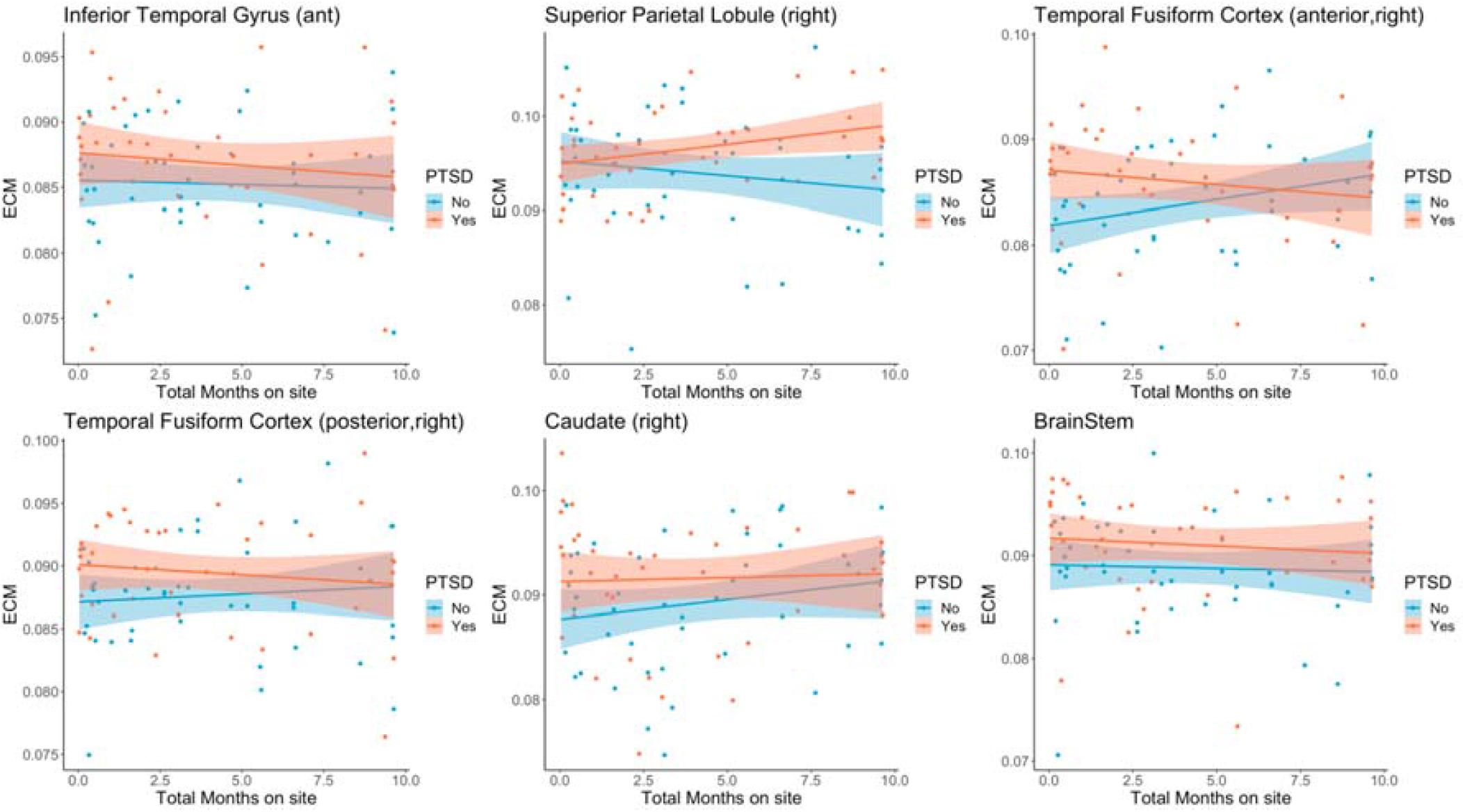
Effects of WTC exposure duration on centrality values for the identified hubs. These panels show the relation between WTC exposure duration expressed in months and eigenvector centrality values for the nine identified hubs. Orange and blue dots represent WTC-responders with and without PTSD respectively.

**Table S2.** Association between EC values, PTSD status and WTC exposure duration. Generalized regression models (GLM) examining WTC exposure duration (i.e., months on site) moderates the association between PTSD (WTC-PTSD vs non-PTSD) and EC values controlling for major depressive disorder (MDD) and medication use (psychotropic and opioid) on eigenvector (EC) value of a single brain area (defined using the Harvard-Oxford atlas).

| Predictors | Inferior Temporal Gyrus (ant) |  |  | Superior Parietal Lobule (right) |  |  | Temporal Fusiform Cortex (anterior,right) |  |  | Temporal Fusiform Cortex (posterior,right) |  |  | Caudate (right) |  |  | BrainStem |  |  |
| --- | --- | --- | --- | --- | --- | --- | --- | --- | --- | --- | --- | --- | --- | --- | --- | --- | --- | --- |
|  | Estimates | CI | p | Estimates | CI | p | Estimates | CI | p | Estimates | CI | p | Estimates | CI | p | Estimates | CI | p |
| Intercept | 0.086 | 0.083 – 0.088 | **<0.001** | 0.095 | 0.092 – 0.098 | **<0.001** | 0.082 | 0.079 – 0.085 | **<0.001** | 0.087 | 0.085 – 0.090 | **<0.001** | 0.088 | 0.085 – 0.091 | **<0.001** | 0.089 | 0.086 – 0.092 | **<0.001** |
| Months on site | -0.000 | -0.001 – 0.000 | 0.841 | -0.000 | -0.001 – 0.000 | 0.296 | 0.001 | 0.000 – 0.001 | **0.042** | 0.000 | -0.000 – 0.001 | 0.591 | 0.000 | -0.000 – 0.001 | 0.161 | -0.000 | -0.001 – 0.000 | 0.821 |
| psychotropic | -0.001 | -0.004 – 0.001 | 0.315 | 0.001 | -0.002 – 0.005 | 0.422 | -0.001 | -0.005 – 0.002 | 0.407 | -0.002 | -0.005 – 0.001 | 0.149 | -0.002 | -0.005 – 0.002 | 0.354 | 0.000 | -0.003 – 0.003 | 0.901 |
| opioid | -0.004 | -0.010 – 0.002 | 0.155 | -0.000 | -0.007 – 0.007 | 0.999 | -0.008 | -0.015 – -0.001 | **0.017** | -0.002 | -0.007 – 0.003 | 0.417 | -0.006 | -0.013 – 0.001 | 0.099 | -0.001 | -0.007 – 0.006 | 0.812 |
| MDD | 0.000 | -0.003 – 0.003 | 0.972 | -0.000 | -0.004 – 0.003 | 0.811 | 0.000 | -0.003 – 0.004 | 0.910 | -0.002 | -0.005 – 0.001 | 0.173 | 0.002 | -0.002 – 0.006 | 0.267 | -0.001 | -0.005 – 0.002 | 0.401 |
| PTSD | 0.002 | -0.001 – 0.006 | 0.196 | -0.000 | -0.004 – 0.004 | 0.901 | 0.005 | 0.001 – 0.009 | **0.008** | 0.004 | 0.001 – 0.007 | **0.015** | 0.003 | -0.001 – 0.007 | 0.163 | 0.003 | -0.001 – 0.007 | 0.105 |
| Months on site*PTSD | -0.000 | -0.001 – 0.001 | 0.862 | 0.001 | -0.000 – 0.001 | 0.105 | -0.001 | -0.001 – 0.000 | 0.061 | -0.000 | -0.001 – 0.000 | 0.639 | -0.000 | -0.001 – 0.001 | 0.533 | -0.000 | -0.001 – 0.001 | 0.832 |
| Observations | 86 |  |  | 86 |  |  | 86 |  |  | 86 |  |  | 86 |  |  | 86 |  |  |
| R<sup>2</sup> | 0.077 |  |  | 0.102 |  |  | 0.156 |  |  | 0.113 |  |  | 0.115 |  |  | 0.061 |  |  |

**Table S3.**
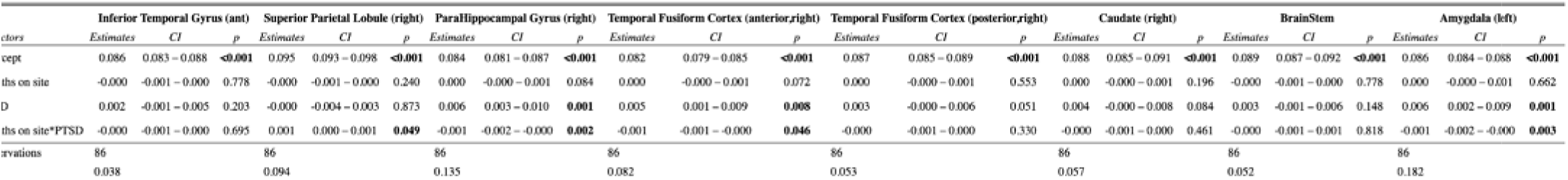
Association between #C values, PTSD status and WTC exposure duration. Series of regression models performed using predictor factors current PTSD diagnosis and, as outcome entered the eigenvector centrality value of a single brain area (defined using the Harvard-Oxford atlas).

